# Improving in silico prediction of influenza vaccine effectiveness by genome analysis incorporating epitope information

**DOI:** 10.1101/2023.02.14.23285900

**Authors:** Lirong Cao, Jingzhi Lou, Qi Li, Hong Zheng, Chris Ka Pun Mok, Zigui Chen, Renee Wan Yi Chan, Peter Pak Hang Cheung, Marc Ka Chun Chong, Eng Kiong Yeoh, William Ka Kei Wu, Jun Yu, Paul Kay Sheung Chan, Benny Chung-Ying Zee, Maggie Haitian Wang

## Abstract

Fast evaluation of vaccine effectiveness (VE) is valuable for facilitating vaccine development and making vaccination strategy. In previous studies, we developed the computational model linking molecular variations and VE for the influenza and COVID-19, through which VE prediction prior to mass vaccination and infection is possible. In this study, we perform a complete survey of the predictive effect of major functional regions of the influenza virus for VE. Interestingly, we found that the genetic distance measured on the antigenic sites being also the effective mutations for epidemics is a strong predictor for influenza VE. Based on the identified optimal predictor codon set, we develop the improved VE-Genetic Distance model for influenza (VE-GD flu). The prediction accuracy of the new model is R-square 87.1% for H3N2 (*p*-value < 0.001) on VE data of the United States. Leave-one-out cross validation shows that the concordance correlation coefficient of the predicted and observed VE is 90.6% (95% CI: 73.8-96.9). Significant prediction improvement is also found for pH1N1. Accurate prediction of influenza VE before vaccine deployment may facilitate reverse vaccinology to optimize vaccine antigen design and government preparedness of influenza epidemics.

## Introduction

Influenza causes numerous morbidity and mortality annually worldwide^1^. Many countries have implemented influenza vaccination programmes to reduce disease burden due to influenza epidemics. However, the performance of influenza vaccines varies by vaccine match to circulating viruses each year. While the protective effect of vaccine can be assessed by vaccine efficacy through randomized control trials (RCTs) or vaccine effectiveness (VE) in observational studies^2-4^, rapid evaluation of VE before mass vaccination can provide timely information for vaccine design and policy-makers to evaluate epidemic risk and design optimal public health strategies.

Previously, we developed a computational framework to predict VE for the influenza^5,6^ and COVID-19^7^ by analysis of virus sequencing data. We found that genetic distance (GD) between vaccine strains and circulating viruses is significantly correlated with vaccine effectiveness (VE). The key to model the relationship between VE and GD lies in the appropriate quantification of GD. We found that GD measured on the effective mutations (EMs) showed good prediction performance of VE, which is composed of mutations significantly associated with epidemic level^8^. Evident linear relationships were identified between VE and GD at the EM sites for influenza A pH1N1, H3N2, and influenza B and were validated in independent data^5^. Given that mutations at antigenic sites or receptor binding domains were biologically responsible for major antigenic drift of the influenza virus^9-11^, we hypothesize that appropriately incorporating their information may further improve the VE prediction on top of the current EM-based model. Therefore, in this study, we conducted a complete survey of genetic mismatch evaluated on the epitope regions on the hemagglutinin (HA) and neuraminidase (NA) proteins and EM sites. We also disentangle the effects of genetic escape at HA and NA on VE through mediation analysis.

The outcome of this study is quantified measures of genetic escape influencing vaccine protection and improved VE prediction model, ready-to-use for early evaluation of influenza VE of a given vaccine strain against any composition of circulating virus population.

## Methods

### Vaccine effectiveness data and viral sequences

The VE data were extracted from published epidemiological studies^12,13^. The VE reports included in the analysis satisfy the following inclusion criteria: (1) the study applied a test-negative case control design or cohort to evaluate VE; (2) used RT-PCR test to confirm influenza infections; (3) the target population is all-age group; (4) the primary outcome is influenza-like illness; (5) the study included vaccine effectiveness against influenza A. Since surveillance of influenza VE in the North America is systematic and continuous, we included all VE observations surveyed in the US and Canada meeting the inclusion criteria. A complete summary of the VE data was described in the supplementary materials **Tables S1-2**.

Virus genetic sequencing data were downloaded from the NCBI^14^ and GISAID EpiFlu databases^15^. All available sequences of influenza A H1N1pdm09 and H3N2 that matched to the flu seasons and locations of the observational studies were extracted, consisted of a total number of 3,974 hemagglutinin (HA) and neuraminidase (NA) protein sequences between 2009/10 and 2018/19 flu seasons. Multiple sequence alignment was performed by MEGA (7.0.26). The influenza vaccine strains recommended by the WHO for use in the Northern Hemisphere were summarized in the Supplement Materials **Table S3**.

### Antigenic sites (AS)

Antigenic sites or epitopes of HA and NA for H1N1 and H3N2 viruses were collected from literature^16-22^, which are identified through experimental approach and resolving virus crystal structure (**Table S4)**.

### Effective mutations (EMs)

The effective mutations (EMs) are a set of mutations that is significantly associated with influenza epidemic levels in a given geographical region. The EM-test was introduced in previous studies with applications on the H3N2 virus^8,23^. Herein, we give a brief description of the method. The EM framework aims to account for the time-dependency of mutation effect for evading population immunity when evaluating their statistical importance. In contrast to the traditional association tests that regard all mutations to have constant effect through time, the EM test simultaneously estimates the relevancy of a genomic position and the effective time period of a mutation. Location-wise, an EM needs to meet the criteria that its mutation prevalence has reached a “dominance threshold” (e.g. 0.5) in population; while temporally, the dominant mutations are also expected to act within its effective mutation period, such that the mutation will not always be accounted as a contributor for escaping population immunity which has already adapted over time.

Let *x*_*jk*_ (*t*) denote the nucleotide or amino acid residue at genome position *j* and time *t. j* ∈*J*, where *J* is full sequence length. Let *p*_*j*_ (*t*) denote mutation prevalence of *x*_*j*_ (*t*). Let 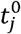 denote the time when *p*_*j*_ (*t*^0^)=0 and *p*_*j*_ (*t*^O^ + 1) > 0. Let θ denote the threshold of mutation prevalence, and 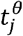 is the earliest time point when *p*_*j*_ (*t*) = θ. The EMs are defined by the following equation,

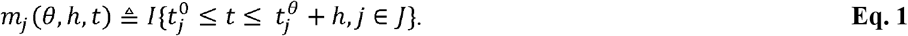

Eq. 1 identifies the EMs with a period, which begins with the first non-zero prevalence of the mutation and ends after *h* intervals following the mutation reaching a prevalence threshold (*θ*. The (*θ, h*) can be estimated by fitting a linear model between the epidemic level y(t) and an aggregated mutation activity metric, the g-measure (*θ, h, t*) = ∑_*j,k*_ *m*_*jk*_ (*θ, h, t*)*p*_*jk*_ (*t*), over a range of (*θ, h*) values, controlling for the covariates including temperate, absolute humidity and season. y(t) in this study is sero-positivity rate of the influenza subtypes by epidemic season, that is, from 1-Oct to 30-Apr. *θ*∈ (0,1) and h = {0,1,2, …}. **Table S5** gives the complete set of fitted values.

The list of EMs identified in the United States can be found in **Table S5**. The AS and EM locations on stereo view of HA and NA protein structures were plotted in **Figures S1-4**, obtained from the RSCB Protein Data Bank (PDB) (https://www.rcsb.org). The summary of AS and the EM sites used in this study is provided in **Figure 1**. These sites serve as candidate codons for exploring their predictive effect for influenza VE. *P*-value <0.01 is considered as statistically significant.

**Figure 1.**
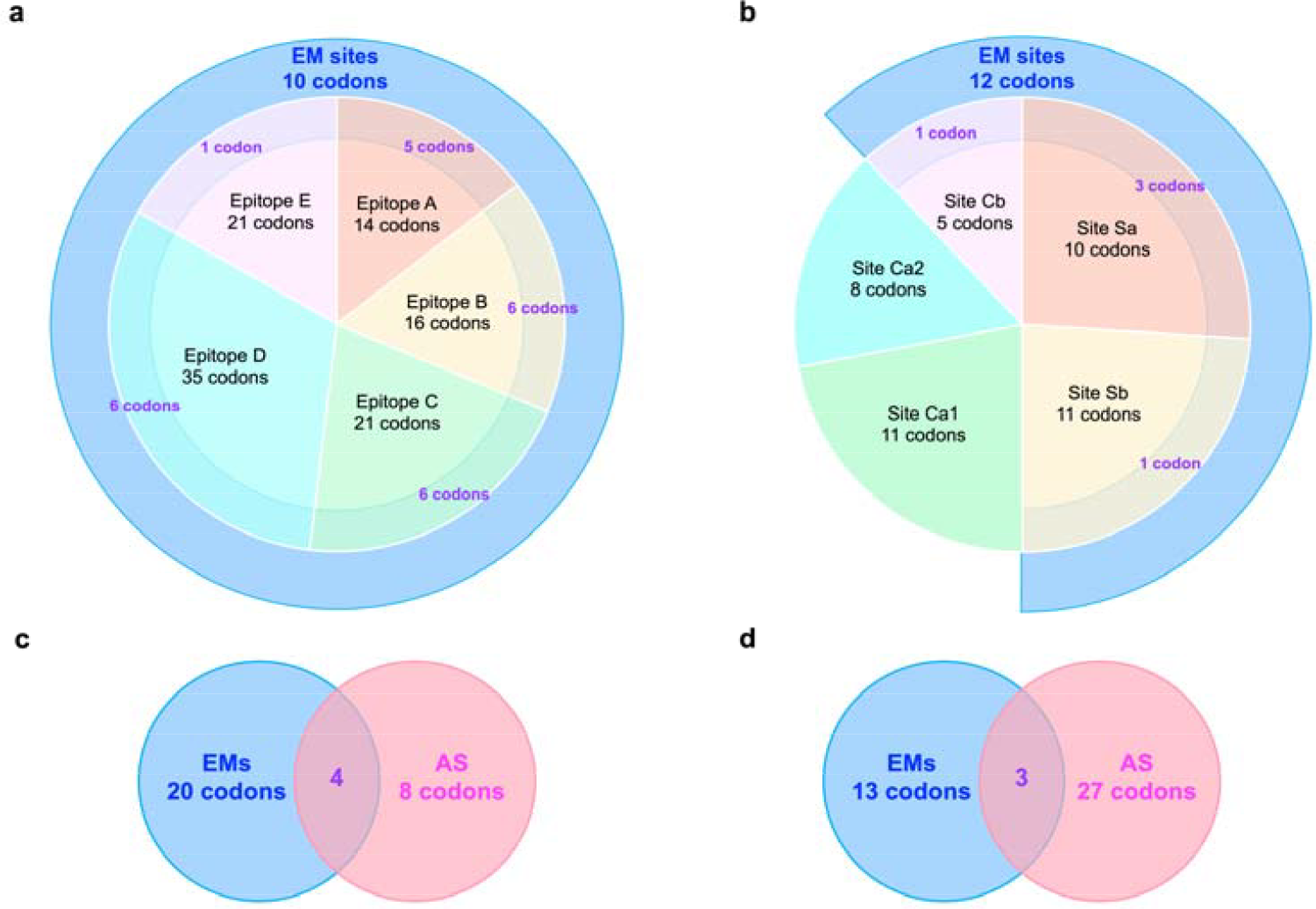
Venn diagram for comparing antigenic sites (AS) and effective mutation (EM) sites. We compared EM sites and AS and identified the intersection codons of two genetic regions. The list of AS and EM sites are provided in supplementary materials Table S4 and Table S5, respectively.

### The VE-GD model for influenza virus (VE-GD flu)

Let W(·) denote a set of predictor codon sites, and *s* denote the protein of concern. *s* = {*HA, NA*}. For a vaccine study *l*, the average genetic mismatch from vaccine strain 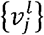 to the circulating viruses on gene *s* corresponding to the study period can be obtained by,

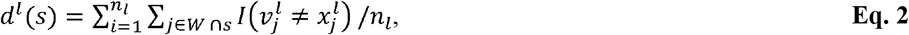

where n_*l*_ is the number of circulating sequences matched to study *l*. To integrate the genetic mismatch from multiple proteins, we utilize the polygenic risk score (PRS) method^24^, which is a widely applied approach in human genetics for disease risk prediction^25,26^. First, the effect of protein s is evaluated by the linear regression,

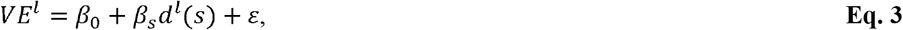

obtaining the estimated coefficient 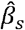. Next, the integrated genetic distance (GD) of HA and NA is obtained by,

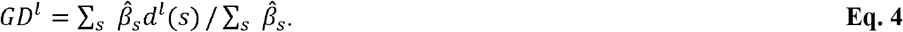

Thus, the VE can be estimated by fitting the linear regression model,

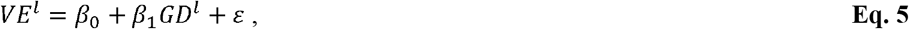

where ε is the random error; *l ∈ L*, and *L* is the total number of VE values. **Eq. 5** captures the relationship between VE and GD for a specific influenza virus subtype. Related VE-GD framework for predicting overall VE involving multiple subtypes is described in Cao et al^5^, and for predicting VE from multiple technology platforms is described in Cao et al^7^.

### Model evaluation

Prediction accuracy is measured by the mean absolute error (MAE) between the predicted and observed VE values, as well as by the concordance correlation coefficient (CCC)^27^. Leave-one-out cross-validation (LOOCV) setting is adopted in which the testing data is independent from the model training data.

### Mediation analysis

With mediation analysis, we investigate the relationship between HA and NA mutations leading to decreased VE. We estimate the effect of HA mutations mediated by the co-mutations occurred on NA protein (referred to as an “indirect effect” in causal mediation terminology), and the drop in VE introduced via HA-only mutations through other immune mechanisms (“direct effect” not mediated by NA mismatch). All analyses was carried out in **R** statistical software^28^ (version 4.0.3) and the R package mediation was used^29^.

## Results

### Relevancy of mutation escape in alternative codon groups for influenza VE by gene

We first evaluate the relevancy of mutation escape in candidate codon sets for VE by gene, and in the following section evaluate the effect of the integrated genetic measure of HA and NA. We found that among all the combinations of epitope sites on the HA and NA and the EMs, the codon set composed of antigenic sites (AS) being also the effective mutations (EMs) is consistently the best estimator for VE of both H3N2 and pH1N1 (*p*-value < 0.01, **Table 1**). Among the five epitopes A-E of HA of H3N2, genetic distance on epitope A and B showed the strongest association among the epitope regions to the reduction of VE for H3N2 (*p*-value <0.01, **Table 1**). This finding is in agreement with the previous understanding of biological roles of these two epitopes towards antigenic drift^30-33^. The EMs are mutations that are found to be significantly associated with epidemic level in the United States during 2003/04-2018/19, which infers potential roles of these mutations in escaping population immunity. Interestingly, we found that when the EMs are not considered, genetic distance on the HA epitopes alone is not predictive to VE for H3N2 (R-sq = 0, *p* = 0.95) and is only weakly associated with VE for pH1N1 (R-sq = 41%, *p* =0.018); however, when genetic distance is evaluated on the AS being EMs, significant relationships are observed (**Fig 2a-b** for H1N1 and **Fig 2d-e** for H3N2). Scatter plots of the relationship of VE and all codon sets can be found in Supplementary Materials **Figures S5-12**.

**Table 1.** Relevancy of genetic escape on alternative codon sets for VE. Legend: *p*-value is significance of the coefficient. ^*^ *p* < 0.01. R-square is goodness-of-fit of Eq. 3 in the training data of the United States.

| VE | Protein | Codon sets | $\hat{\beta}_s$ (SE) | R<sup>2</sup> | p-value |
| --- | --- | --- | --- | --- | --- |
| pH1N1 | HA | All Antigenic Sites (AS) | -7 (2.5) | 41.0% | 0.018 |
|  |  | Site Sa | -11.2 (7.1) | 18.5% | 0.142 |
|  |  | Site Sb | 2.1 (8.4) | 0.6% | 0.808 |
|  |  | Site Ca1 | -3.9 (6.1) | 3.6% | 0.534 |
|  |  | Site Ca2 | -0.5 (28.5) | 0 | 0.986 |
|  |  | Site Cb | -4.3 (9.3) | 1.9% | 0.653 |
|  |  | AS excluding EM | -1.2 (6.3) | 0.4% | 0.847 |
|  |  | EM | -2.3 (0.8) | 41.7% | 0.017 |
|  |  | **EM-AS** | **-8.4 (2.7)** | **47.3%** | **0.009\*** |
|  | NA | AS | -9.9 (2.9) | 51.7% | 0.006* |
|  |  | AS excluding EM | -16.5 (22.5) | 4.6% | 0.479 |
|  |  | EM | -2.2 (0.7) | 51.2% | 0.006 |
|  |  | **EM-AS** | **-10.6 (3.1)** | **52.4%** | **0.005\*** |
| H3N2 | HA | All Antigenic Sites (AS) | -0.1 (2.0) | 0 | 0.953 |
|  |  | Epitope A | -28.6 (8.5) | 48.5% | 0.006* |
|  |  | Epitope B | -9.1 (2.2) | 57.7% | 0.002* |
|  |  | Epitope C | 14.3 (6.3) | 30.1% | 0.042 |
|  |  | Epitope D | 7.3 (3.9) | 22.8% | 0.084 |
|  |  | Epitope E | 8.9 (5.2) | 19.4% | 0.116 |
|  |  | AS on epitope A and B | -7.7 (1.7) | 61.5% | < 0.001* |
|  |  | AS excluding EM | -11 (5.8) | 23.0% | 0.08 |
|  |  | EM | -6.7 (2.0) | 49.8% | 0.005* |
|  |  | **EM-AS** | **-11 (2.3)** | **65.1%** | **< 0.001\*** |
|  | NA | AS | -19.8 (2.7) | 81.6% | < 0.001* |
|  |  | AS excluding EM | 47.1 (75.3) | 3.1% | 0.543 |
|  |  | EM | 1.1 (2.5) | 1.6% | 0.667 |
|  |  | **EM-AS** | **-20.2 (2.7)** | **81.8%** | **< 0.001\*** |

**Figure 2.**
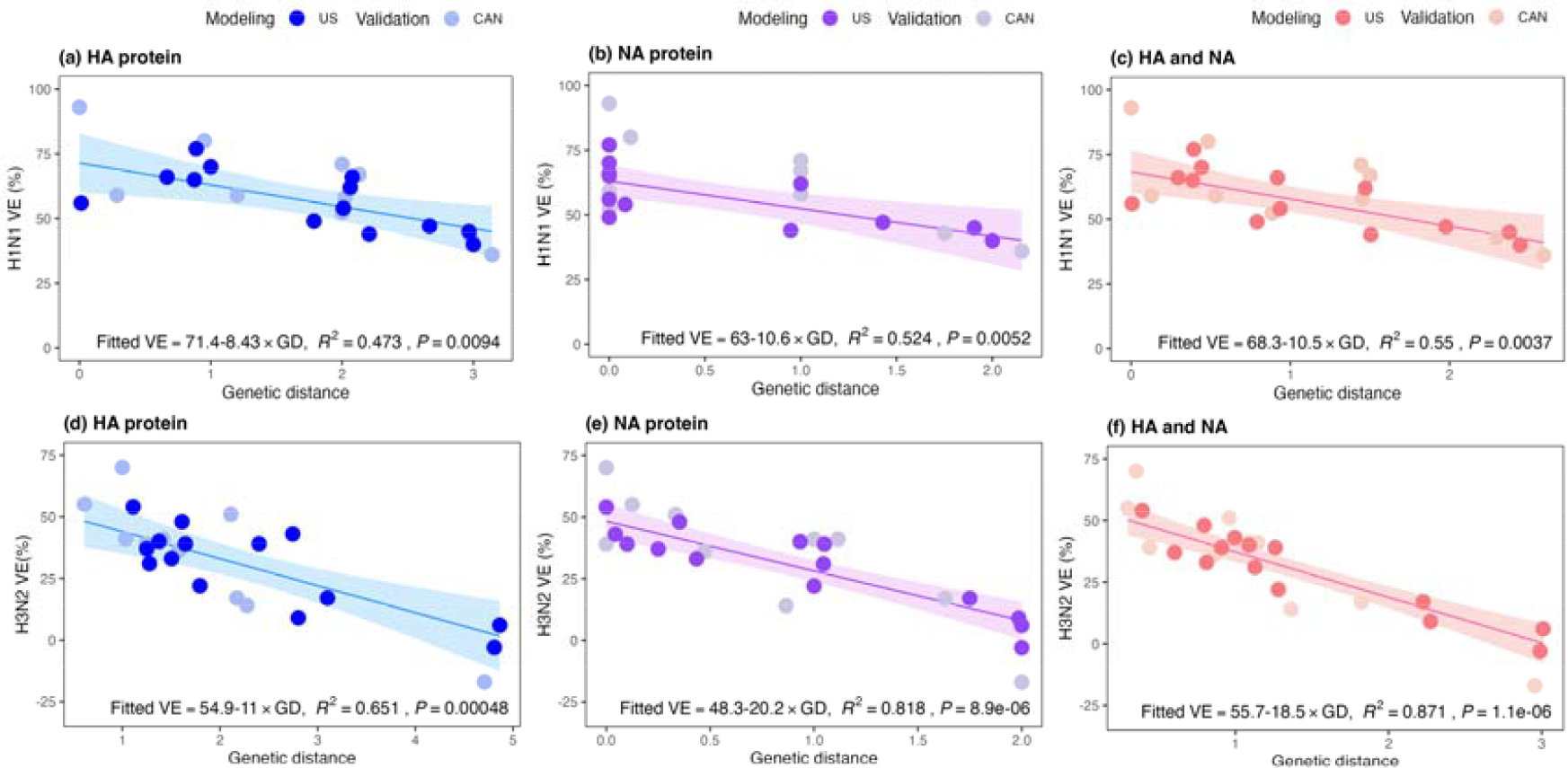
Relationship between vaccine effectiveness (VE) and genetic distance (GD) on the antigenic sites (AS) being also effective mutations (EMs). Legend: Evaluation the relationship between VE and GD calculated on the best predictor codon set, EM-AS. Panels show the analysis by gene: (a) HA of H1N1, (b) NA of H1N1, (c) HA and NA of H1N1; (d) HA of H3N2, (d) NA of H3N2, and (e) HA and NA of H3N2. Dark dot: training data of the United States; light dot: validation data of Canada

### Relationship of influenza VE and the integrative genetic distance of HA and NA

Next, we estimate the relationship of VE and the integrated genetic distance (GD) of HA and NA using the predictor codon set composed of EM-AS. We found that the GD explains 87.1% (*p* < 0.001, **Fig 2c**) variations of VE for the H3N2 and 55% (*p* = 0.0037, **Fig 2f**) of the variations of VE for pH1N1. For each genetic mismatch occurred in the EM-AS loci, the expected VE reduction is 18.5% [95% CI: 14.4-22.5] for the H3N2 and 10.5% [95% CI:4.9-16.1] for pH1N1. Therefore, per mutation impact on VE for the H3N2 is 1.8-fold more than that of pH1N1, indicating the particular challenge of vaccine strain selection for the H3N2 virus. In the ideal situation when a perfect vaccine match is achieved, the expected VE for the H3N2 is 55.7%, lower than the expected VE 68.3% for the pH1N1. The ceiling of VE of the two subtypes might be explained by the higher genetic diversity of the H3N2 virus comparing to the H1N1^34,35^, such that it is more difficult to identify a single presentative strain for the entire virus population of the H3N2.

### Model validation by independent data

First, leave-one-out cross validation was conducted for the VE-GD flu model. The average distance between the observed values and the predicted VE was 7.1% for H1N1 and 6.3% for H3N2. The calibration plot of **Figure 3** depicts that the validation data points are surrounding the 45° line, meaning that the predictions are nearly identical to the observed VE. The test statistic CCC was 90.6% (95% CI: 73.8-96.9) for the H3N2 model and 60.2% (95% CI: 13.3-85.1) for the H1N1 model. In **Figure 2**, the independent data from Canada also show an obvious negative relationship between GD and VE against pH1N1 and H3N2, closely clustering around the fitted line of US models for both single protein and two protein models. In addition, we conducted retroactive predictions for 2019/20 to 2022/23 flu seasons (**Figure 4**, supplementary materials **Table S6)** and found that the predicted H3N2 VE for US was 33.5% (95% prediction interval: 19.2-47.8), close to the observed value of 36% (95% CI: 20-49) in 2021/22 flu seasons^36^. In the next flu season, the estimated H3N2 VE was 49.0% (95% prediction interval: 34.0-64.1), which is in close agreement with the reported effectiveness of 45% (95% CI: 29-58) by US CDC^37^, indicating good predictive power of the model.

**Figure 3.**
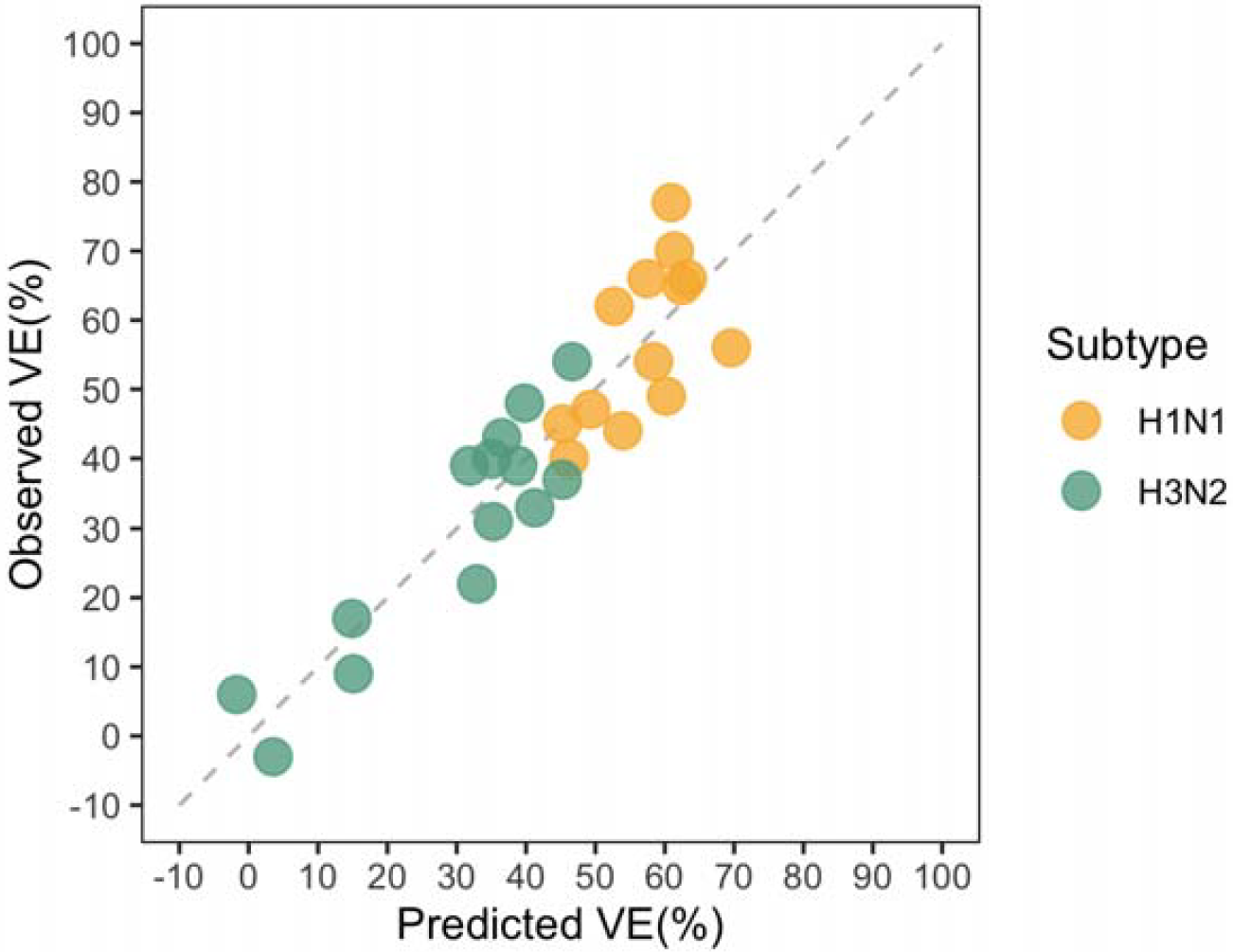
Calibration plot for the prediction accuracy in leave-one-out cross validation. Data points near the dashed line mean that the predicted value is close to the observed VE.

**Figure 4.**
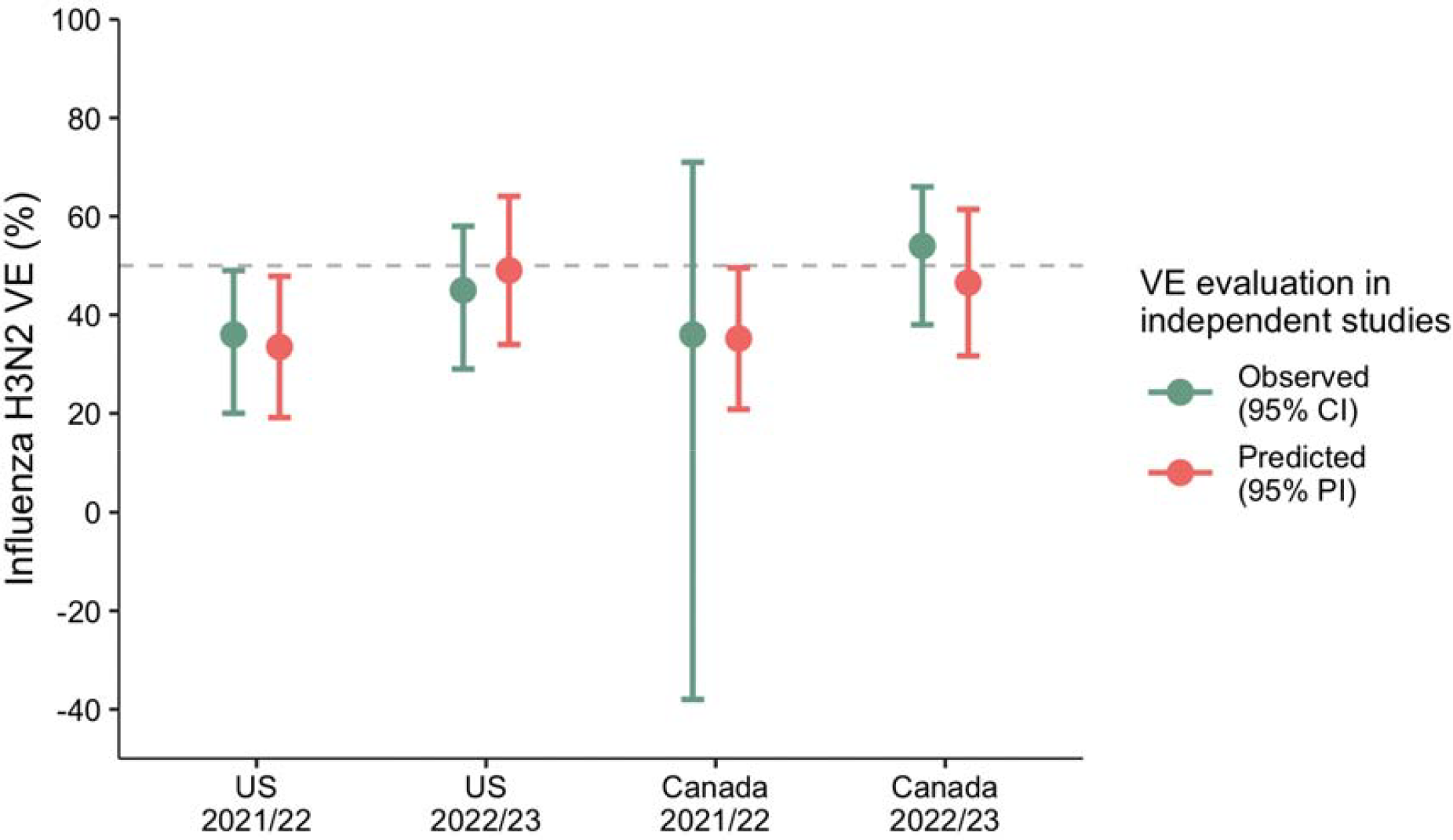
Retroactive prediction for H3N2 VE from 2019/20 to 2022/23 flu seasons. VEs from 2019/20 to 2022/23 flu seasons were retroactively predicted.

### Mediation analysis of genetic factors on vaccine effectiveness

Using the mediation analysis, we estimated that the average direct effect (ADE) of HA excluding the effect of NA is 4.7% (95% CI: 1.5 – 8.2, *p* =0.01) decrease in VE for H3N2 per mismatch; and the indirect effect of HA mediated through the NA is 6.3% (95% CI: 1.5 – 10.6, *p* =0.02) drop in VE per mismatch (**Figure 5**). The indirect effect due to HA-NA mismatch pathway accounts for 57% of the total effect of HA on VE. This result indicates the importance of considering HA and NA jointly in the optimization of influenza vaccine antigen for the H3N2. Currently, no significant indirect effect has been detected for pH1N1 virus.

**Figure 5.**
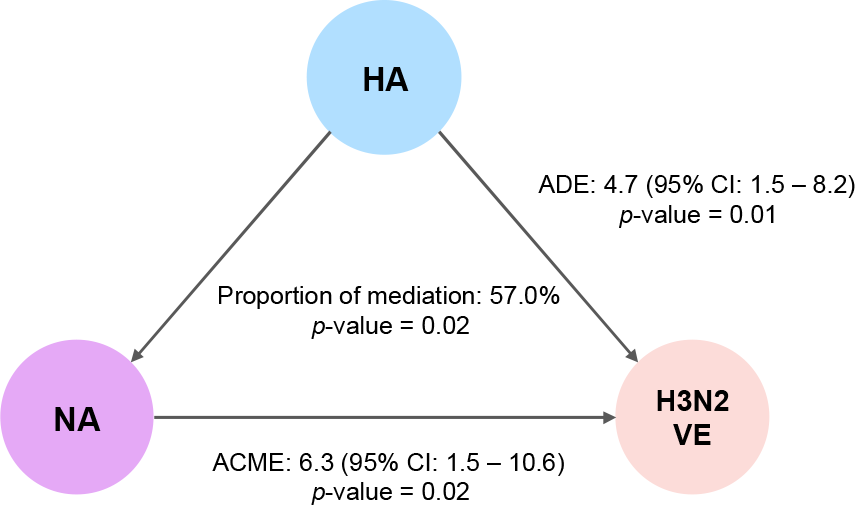
Mediation analysis of the effect of HA and NA on the VE of influenza H3N2. ADE: average direct effect; ACME: average causal mediation effects.

## Discussion

The use of GD to predict vaccine protection can help to evaluate the impact of influenza immunization programme before mass vaccination and inform vaccination policy details, such as determining whether to introduce a particular vaccine in a specific population or identifying preferred vaccine products for tropical or subtropical regions. In this study, we optimized GD measure for HA and NA by feature selection and found that GD on the intersection set of EM and AS is highly predictive of VE of influenza H1N1 and H3N2. However, the remaining AS but not EM sites showed few potentials in VE evaluation. The genetic mismatch on these AS but non-EM sites of HA and NA was consistently uncorrelated with VE of both influenza A subtypes. While literature reported amino acid substitutions emerged on HA or NA may weaken vaccine protection^38,39^, few studies can quantify their effect on VE. This study bridges the molecular-level variations with population-level immunity and refines the quantification method of genetic distance in order to *in silico* predict the effectiveness of influenza vaccines in real time.

This study demonstrated that immunodominant antigenic epitopes are the optimal target to measure GD. Previous research reported that residues in epitopes A and B on HA of influenza H3N2 viruses are important in binding antibodies and genetic variants in these epitopes can help escape antibody response^30-33^. In line with previous findings, our results showed that genetic mismatch on epitopes A and B is associated with VE as amino acid changes on these epitopes could result in vaccine-mismatched influenza viruses predominant thereby causing a reduction in vaccine protection. Similar results are found for the H1N1pdm09 virus that the combined mismatch of sites Sa and Sb on HA can link to VE. The research on selective pressure also demonstrated that positively selected codons were associated with antibody combining epitopes A and B, while not all codons in these sites had predictive value for the evolution of human influenza A and only a few codons on epitopes C, D and E were under positive selection^40^, which also support our findings.

Our previous research developed a novel bioinformatics approach to identify key residues relevant to escape from herd immunity, namely EM sites, and demonstrated the feasibility of using these sites to assess VE but failed for NA of H3N2. This work successfully determined genetic measure for H3N2 NA. While genetic mismatch on AS of NA showed the best performance, the superiority compared to using EM-AS codons is relatively small with a 9.1% difference in terms of the goodness of fit, which also proves the importance of EM-AS in VE evaluation. Previous research also tried to use the dominant epitope as that which shows the most antigenic drift to estimate VE^41-43^. However, a precise determination of which epitopes are dominant was not completed. This study provides useful evidence for identifying key epitopes and measuring vaccine matching levels.

This work can also inform ongoing research efforts to understand the role of NA in the effect of HA mutations on VE. The existing evidence showed that NA-induced antibodies could also provide protection against influenza viruses^44,45^ and mutations on NA can also affect viral antigenicity^46^. Inclusion of HA and NA in influenza vaccines provides enhanced protection against influenza-associated illness compared to HA alone^47-49^. From the perspective of genetic epidemiology, we uncovered the potential mechanisms underlying the association of HA mismatch with VE that a significant mediating effect of NA mismatch existed for H3N2.

The limitations of this study include that only a few countries annually investigate the effectiveness of influenza VE and thus it is still a challenge to customize a prediction model for each region. Moreover, numerous studies reported that pre-existing immunity from previous infection and vaccinations can also affect vaccine protective effects^50-55^. The antigenic distance (AD) hypothesis proposed that when prior and current vaccines are antigenically distinct, the influence of the former vaccination should be minimal^56^. When their AD is smaller, positive interference to the current vaccine protection is expected when the AD between prior vaccines and epidemic strains is small, while the interference becomes negative when the AD between pre-existing immunity and circulating viruses is large. In future studies, we will attempt to quantify the impact of pre-existing immunity on VE as data on immune history increases. Despite the limitations, the relationship between GD and VE is deterministic and the key mutation positions involved in this study are also well demonstrated in plenty of biology experiments and bioinformatics analyses. Improved quantification of GD based on EM-AS codons could assist real-time prediction of VE and candidate vaccine strain selection. The predictive models for VE developed herein may facilitate future vaccine deployment and resource coordination.

## Data Availability

All data produced in the present study are available upon reasonable request to the authors.

## Acknowledgements

We thank the contributions of all the health care workers and scientists, the GISAID team, and the submitting and the originating laboratories. This work was partially supported by the Strategic Seed Funding for Collaborative Research Scheme [3136001], the Chinese University of Hong Kong Direct Grant [2022.02], and NSFC Excellent Young Scientists Fund [32322088]. The funders had no role in study design, data collection and analysis, decision to publish or preparation of the manuscript.

## Author Contributions

M.H.W conceived the study. L.C processed data, carried out the analysis and wrote the manuscript. J.L., Q.L., H.Z., C.K.P.M, Z.C., R.W.Y.C., P.P.H.C., M.K.C.C., E.K.Y., P.K.S.C., and B.C.Y.Z. critically read and revised the manuscript and gave final approval for publication.

