## Supplementary materials for "Improving in silico prediction of influenza vaccine effectiveness by genome analysis incorporating epitope information"

### S1 Supplementary tables

#### Table S1. Influenza vaccine effectiveness in United States, 2009-2019

| Flu season | H1N1 VE (%) | H3N2 VE (%) | Source |
| --- | --- | --- | --- |
| 2009-10 | 56 | - | US CDC^1^ |
| 2010-11 | 66 | 54 | US CDC^2^ |
| 2011-12 | 65 | 39 | US CDC^3^ |
| 2012-13 | - | 39 | US CDC^4^ |
| 2013-14 | 54 | 17 | US CDC^5^ |
| 2014-15 | - | 6 | US CDC^6^ |
| 2015-16 | 45 | 43 | US CDC^7^ |
| 2016-17 | - | 33 | US CDC^8^ |
| 2017-18 | 62 | 22 | US CDC^9^ |
| 2018-19 | 44 | 9 | US CDC^10^ |
| 2010-11 | 70 | 37 | Ohmit SE 2013^11^ |
| 2012-13 | - | 40 | Ohmit SE 2015^12^ |
| 2013-14 | 66 | - | Ohmit SE 2016^13^ |
| 2014-15 | - | −3 | Petrie JG 2017^14^ |
| 2010-11 | 77 | 48 | Bateman AC 2013^15^ |
| 2012-13 | - | 31 | McLean HQ 2014^16^ |
| Mean | 60.50 | 29.64 | - |
| Standard Deviation | 10.65 | 16.93 | - |
| Coefficient of Variation | 17.6% | 57.1% | - |

#### Table S2. Influenza vaccine effectiveness in Canada, 2009-2019

| Flu Season | H1N1 VE (%) | H3N2 VE (%) | Source |
| --- | --- | --- | --- |
| 2005-06 | - | 70 | BC CDC^17^ |
| 2006-07 | - | 41 | BC CDC^18^ |
| 2007-08 | - | 57 | BC CDC^19^ |
| 2008-09 | - | 55 | BC CDC^20^ |
| 2009-10 | 93 | - | BC CDC^21^ |
| 2010-11 | 59 | 39 | BC CDC^22^ |
| 2011-12 | 80 | 51 | BC CDC^23^ |
| 2012-13 | 59 | 41 | BC CDC^24^ |
| 2013-14 | 71 | - | BC CDC^25^ |
| 2014-15 | - | -17 | BC CDC^26^ |
| 2015-16 | 43 | - | BC CDC^27^ |
| 2016-17 | - | 36 | BC CDC^28^ |
| 2017-18 | 58 | 14 | BC CDC^28^ |
| 2018-19 | 67 | 17 | BC CDC^29^ |
| Mean | 66·25 | 36.73 | - |
| Standard Deviation | 15·31 | 24.27 | - |
| Coefficient of Variation | 23·1% | 66.1% | - |

#### Table S3. WHO recommended vaccine strains for Northern hemisphere, 2009-2023

| Flu season | H1N1pdm09 | H3N2 |
| --- | --- | --- |
| 2009-10 | A/California/7/2009 | - |
| 2010-11 | A/California/7/2009 | A/Perth/16/2009 |
| 2011-12 | A/California/7/2009 | A/Perth/16/2009 |
| 2012-13 | A/California/7/2009 | A/Victoria/361/2011 |
| 2013-14 | A/California/7/2009 | A/Victoria/361/2011 |
| 2014-15 | A/California/7/2009 | A/Texas/50/2012 |
| 2015-16 | A/California/7/2009 | A/Switzerland/9715293/2013 |
| 2016-17 | A/California/7/2009 | A/Hong Kong/4801/2014 |
| 2017-18 | A/Michigan/45/2015 | A/Hong Kong/4801/2014 |
| 2018-19 | A/Michigan/45/2015 | A/Singapore/INFIMH-16-0019/2016 |
| 2019-20 | A/Brisbane/02/2018 | A/Kansas/14/2017 |
| 2020-21 | A/Guangdong-Maonan/SWL1536/2019 | A/Hong Kong/2671/2019 |
| 2021-22 | A/Victoria/2570/2019 | A/Cambodia/e0826360/2020 |
| 2022-23 | A/Victoria/2570/2019 | A/Darwin/6/2021 |

#### Table S4. List of antigenic sites (AS)

| **Influenza A** | **Genes** | **Epitopes** | **Amino acid position** |
| --- | --- | --- | --- |
| H3N2 | HA gene^30-32^  (H3 numbering) | Epitope A | 122,124,126,130,131,132,133,135,137,138,140,142,143,144,145,146,150,152,168 |
|  |  | Epitope B | 128,129,155,156,157,158,159,160,163,164,165,186,187,188,189,190,192,193,194,  196,197,198 |
|  |  | Epitope C | 44,45,46,47,48,50,51,53,54,273,275,276,278,279,280,294,297,299,300,304,305,307,  308,309,310,311,312 |
|  |  | Epitope D | 96,102,103,117,121,167,170,171,172,173,174,175,176,177,179,182,201,203,205,207,  208,209,212,213,214,215,216,217,218,219,226,227,228,229,230,240,242,244,246,247,  248 |
|  |  | Epitope E | 57,59,62,63,67,75,78,80,81,82,83,86,87,88,91,92,94,109,260,261,262,265 |
|  | NA gene^33^ | | 150,198,199,220,221,253,329,334,344,368,370,403 |
| H1N1 | HA gene^34,35^  (H3 numbering) | Site Sa | 128,129,156,157,158,159,160,162,163,164,165,166,167 |
|  |  | Site Sb | 187,188,189,190,191,192,193,194,195,196,197,198 |
|  |  | Site Ca1 | 169,170,171,172,173,206,207,208,238,239,240 |
|  |  | Site Ca2 | 140,141,142,143,144,145,224,225 |
|  |  | Site Cb | 78,79,81,82,83,122 |
|  | NA gene^36^ | | 93,94,95,216,217,219,220,221,250,251,252,254,262,263,264,265,266,267,268,270,  355,358,375,377,378,388,389,449,450,451 |

#### Table S5. List of effective mutation (EM)^37^ sites for human influenza A viruses

| **Flu type** | **Protein** | **(*θ, h*)** | ***p*-value** | **Total number** | **EM sites list** |
| --- | --- | --- | --- | --- | --- |
| H1N1pdm09 | HA | (0.8, 0) | 0.001 | 17 | 13, 91, 101, 114, 160, 179, 180, 181, 200, 202, 214, 233, 273, 300, 312, 391, 468 |
| H1N1pdm09 | NA | (0.8, 0) | 0.004 | 16 | 13, 34, 40, 44, 77, 81, 106, 188, 200, 264, 270, 314, 321, 386, 432, 449 |
| H3N2 | HA | (0.6, 0) | 0.036 | 34 | 19, 49, 61, 64, 66, 78, 137, 147, 156, 158, 160, 161, 174, 175, 176, 187, 189, 205, 209, 214, 228, 229, 239, 241, 277, 294, 327, 328, 377, 391, 422, 466, 500, 505 |
| H3N2 | NA | (0.9, 5) | 0.13 | 24 | 81, 93, 147, 150, 176, 194, 215, 221, 245, 247, 267, 310, 329, 339, 344, 367, 369, 370, 372, 380, 387, 402, 464, 468 |

Note that *θ* and *h* were estimated by optimizing the fitness function of the g-measure and some covariates including mean temperature, absolute humidity and season against annual sero-positivity rate. The annual mean temperature (centigrade) and mean absolute humidity (g/m^3^) were retrieved from Weather Underground via <https://www.wunderground.com/>. The yearly number of positive cases and specimens were retrieved from New York Department of Health (NYDOH) via <https://www.health.ny.gov/diseases/communicable/influenza/surveillance/>. The annual sero-positivity rate is calculated as number of positive cases divided by number of specimens.

#### Table S6. Prospective prediction performance for H3N2 from 2019/20 to 2022/23 flu seasons

| **No.** | **Country** | **Flu season** | **Observed VE**  **(95% CI)** | **Predicted VE**  **(95% prediction interval)** | **References** |
| --- | --- | --- | --- | --- | --- |
| [1] | US | 2021/22 | 36 (20-49) | 33.5 (19.2-47.8) | US CDC^38^ |
| [2] | US | 2022/23 | 45 (29-58) | 49.0 (34.0-64.1) | US CDC^39^ |
| [3] | Canada | 2021/22 | 36 (−38-71) | 35.2 (22.8-50.0) | BC CDC^40^ |
| [4] | Canada | 2022/23 | 45 (29-58) | 46.6 (31.7-61.4) | BC CDC^41^ |

### S2 Supplementary figures

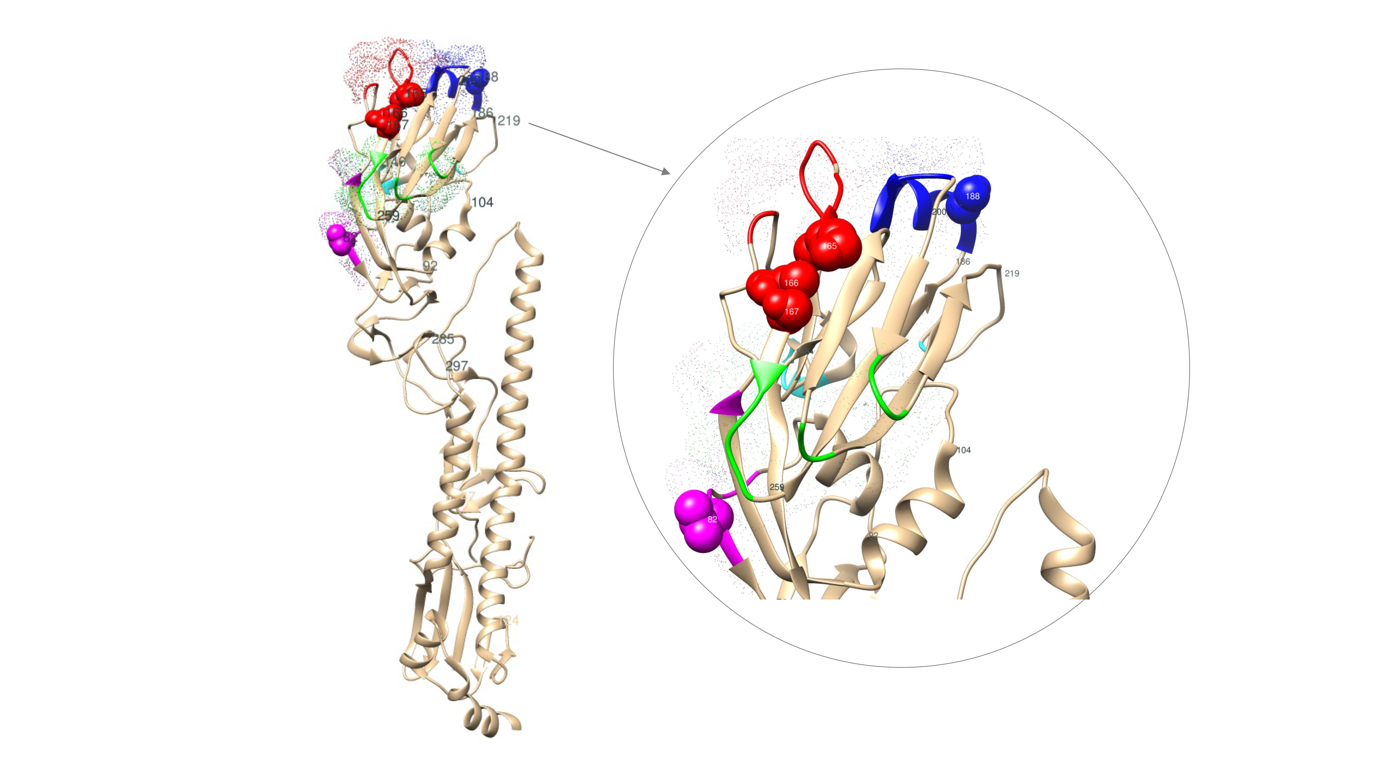

#### Figure S1 Stereo view showing the intersection sets of EM sites and AS on HA protein of H1N1

The intersection sets of EM sites and AS (colored sphere) are displayed on 3D structure of HA protein. The codons with colored dot shadow are five antigenic regions reported by literature^34,35^. The codons marked with positions are EM sites proposed in our previous study^37^. Chimera was used to generate the stereo view of HA protein structure with 5W5U^42^ of Protein Data Bank.

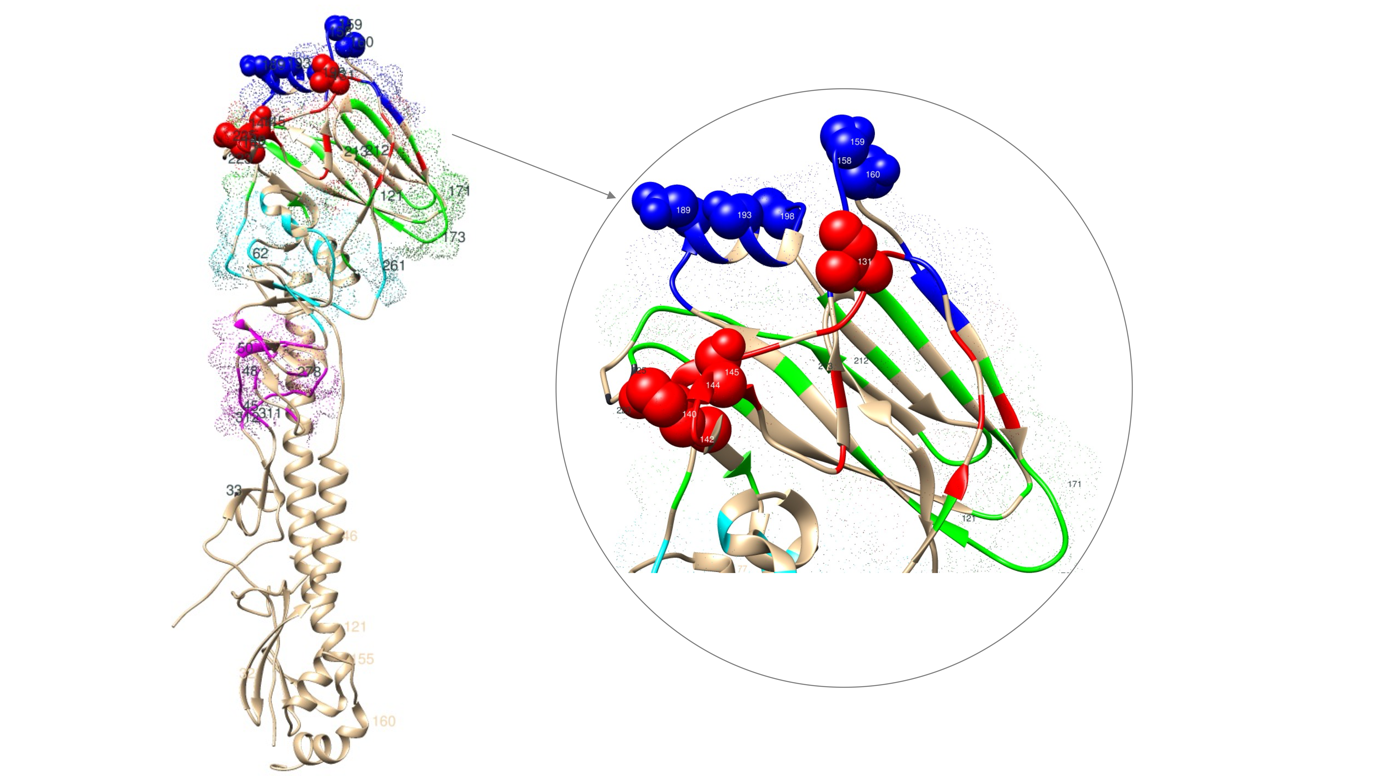

#### Figure S1 Stereo view showing the intersection sets of EM sites and AS on HA protein of H3N2

The intersection sets of EM sites and AS (colored sphere) are displayed on 3D structure of HA protein. The codons with colored dot shadow are five epitopes reported by literature^30-32^. The codons marked with positions are EM sites proposed in our previous study^37^. Chimera^43^ was used to generate the stereo view of HA protein structure with 2VIU^44^ of Protein Data Bank. The structural analysis was restricted to codons annotated in the PDB structure file.

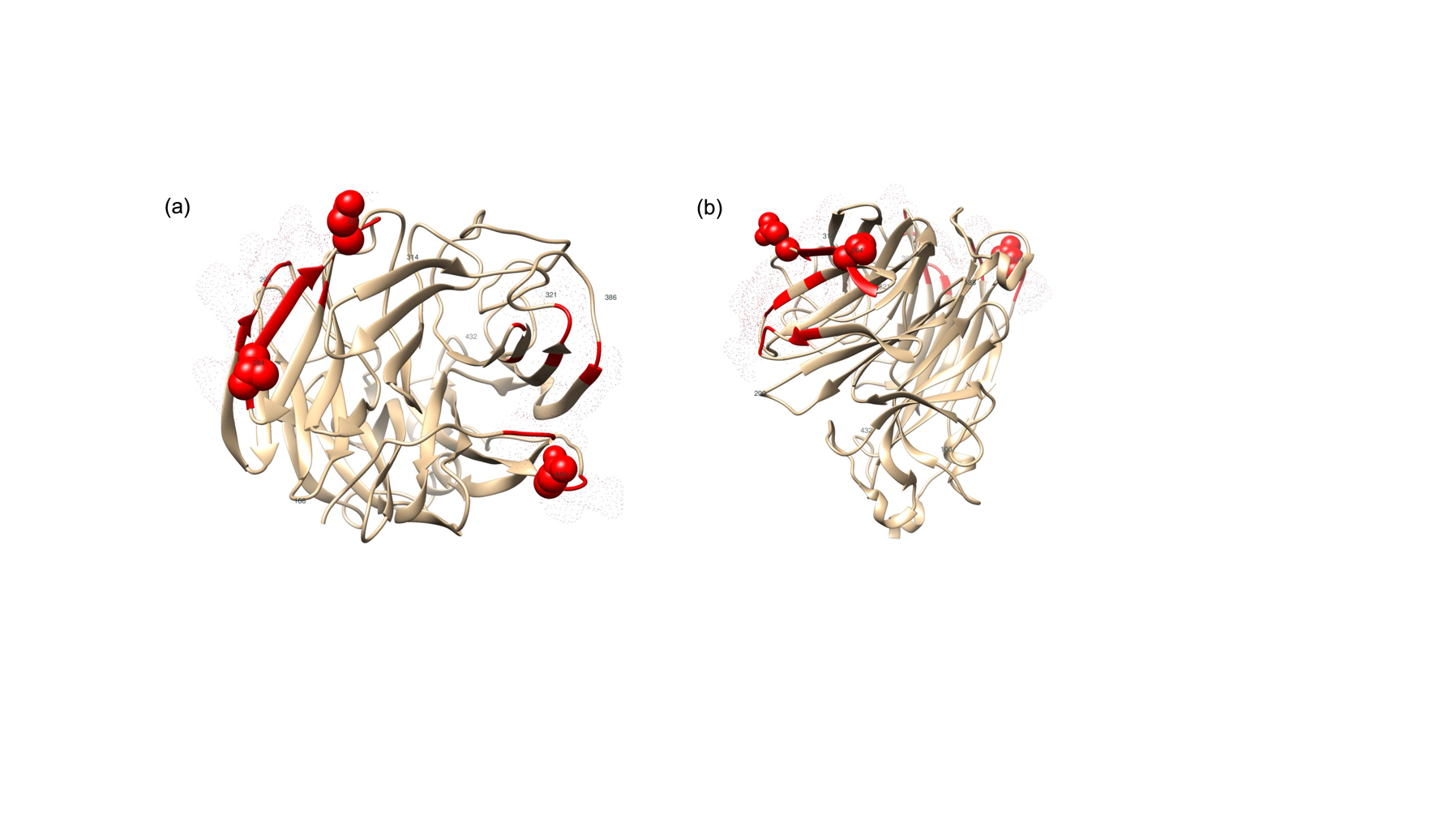

#### Figure S3 Stereo view showing the intersection sets of EM sites and AS on NA protein of H1N1

The intersection sets of EM sites and antigenic sites (red sphere) are displayed on 3D structure of NA protein. The codons with red dot shadow are antigenic sites identified by Wan et al^36^. The codons marked with positions are EM sites proposed in our previous study^37^. Chimera was used to generate the stereo view of NA protein structure with 4B7Q^45^ of Protein Data Bank. Panel (a) is top view, normal to the viral membrane and shows EM-AS sites on the globular head of the NA. Panel (b) is side view, parallel to the viral membrane.

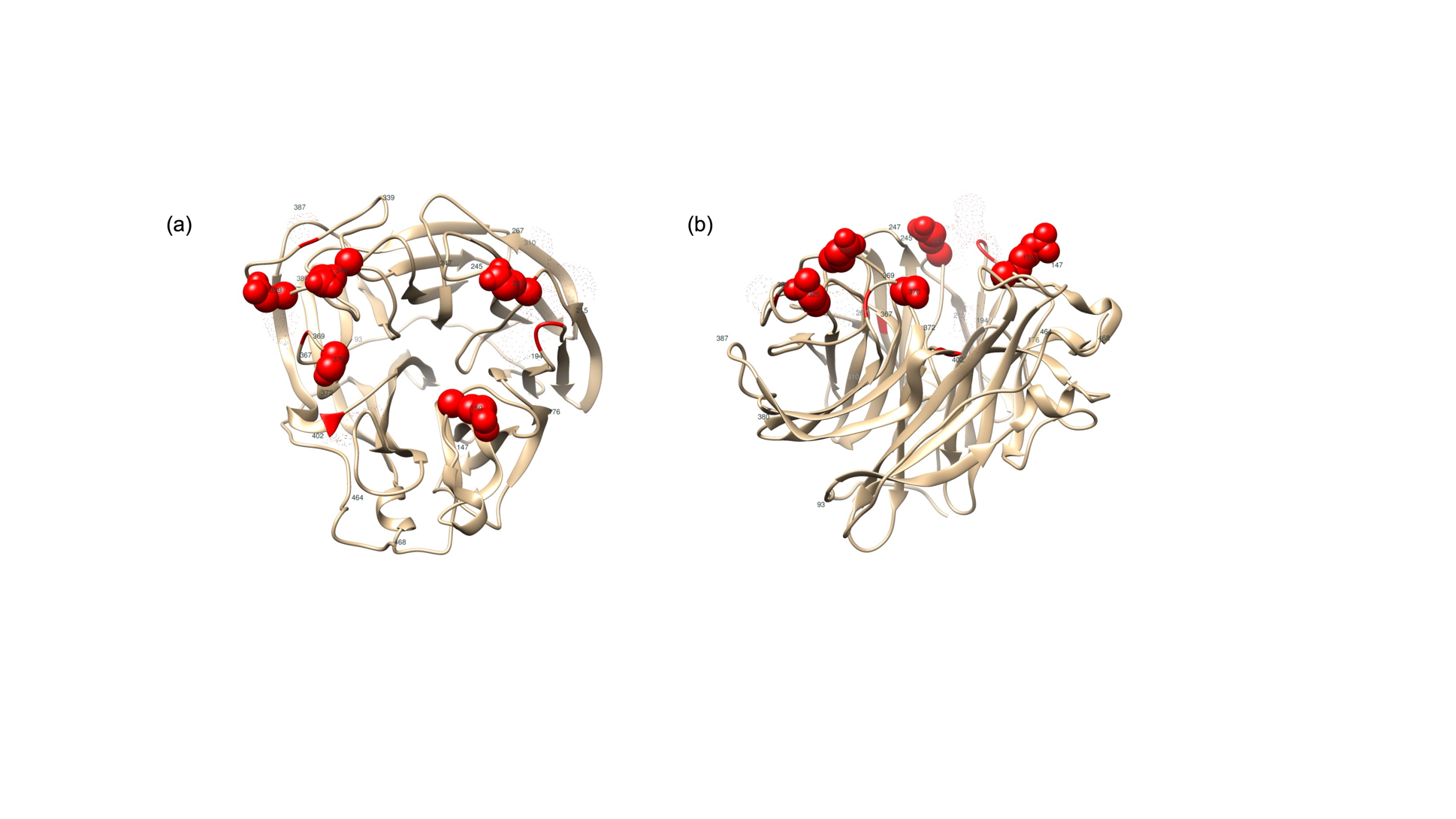

#### Figure S4 Stereo view showing the intersection sets of EM sites and AS on NA protein of H3N2

The intersection sets of EM sites and AS (red sphere) are displayed on 3D structure of NA protein. The codons with red dot shadow are antigenic sites identified by Gulati et al^33^. The codons marked with positions are EM sites proposed in our previous study^37^. Chimera was used to generate the stereo view of NA protein structure with 4GZQ^46^ of Protein Data Bank. Panel (a) is top view, normal to the viral membrane and shows EM-AS sites on the globular head of the NA. Panel (b) is side view, parallel to the viral membrane.

### S3 Supplementary results

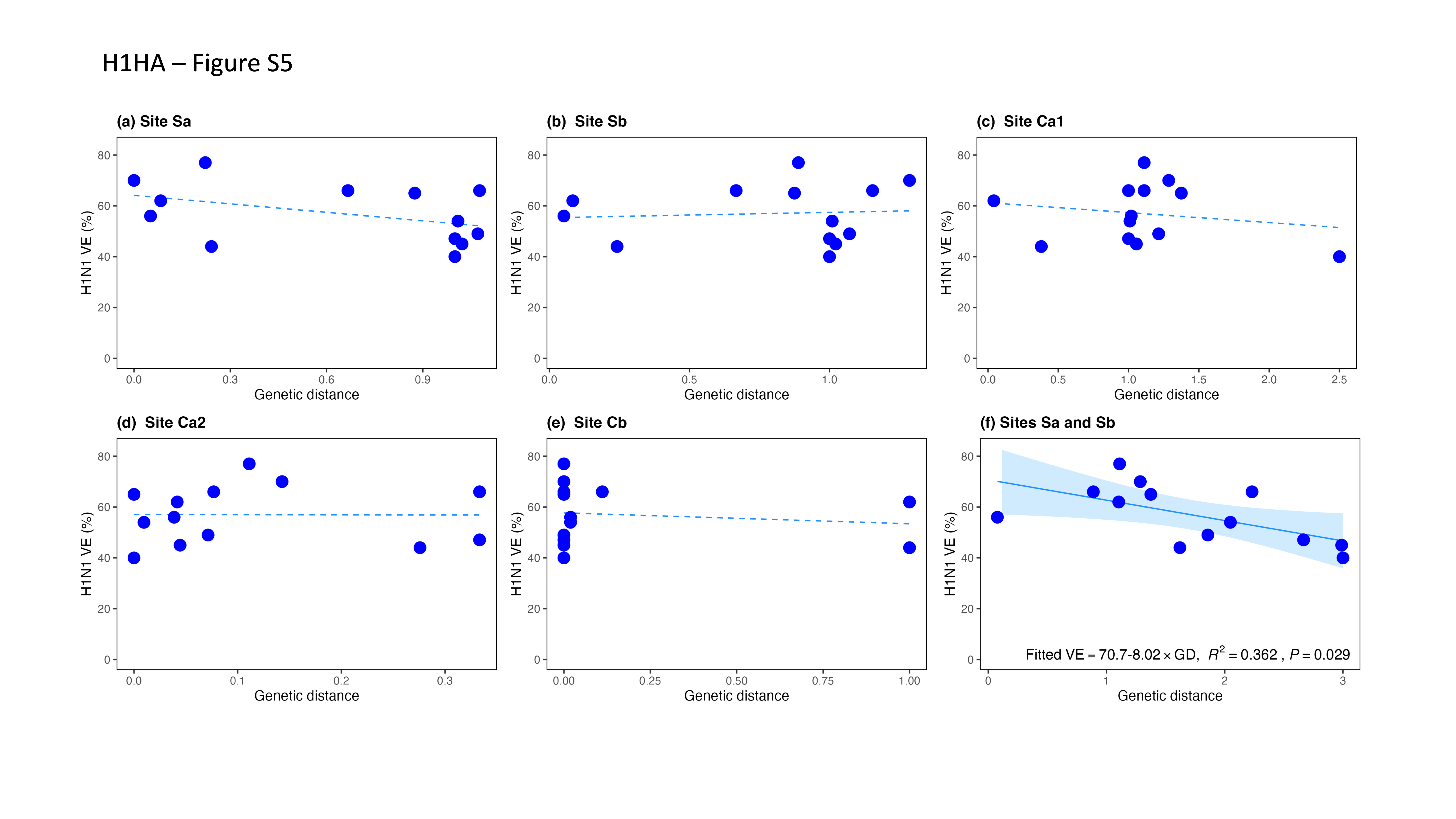

#### Figure S5. The relationship between H1N1 VE and genetic distance (GD) on five AS of HA protein

(a) H1N1 VE against GD on site Sa; (b) H1N1 VE against GD on site Sb; (c) H1N1 VE against GD on site Ca1; (d) H1N1 VE against GD on site Ca2; (e) H1N1 VE against GD on site Cb; (f) H1N1 VE against the integrated GD of Sa and Sb.

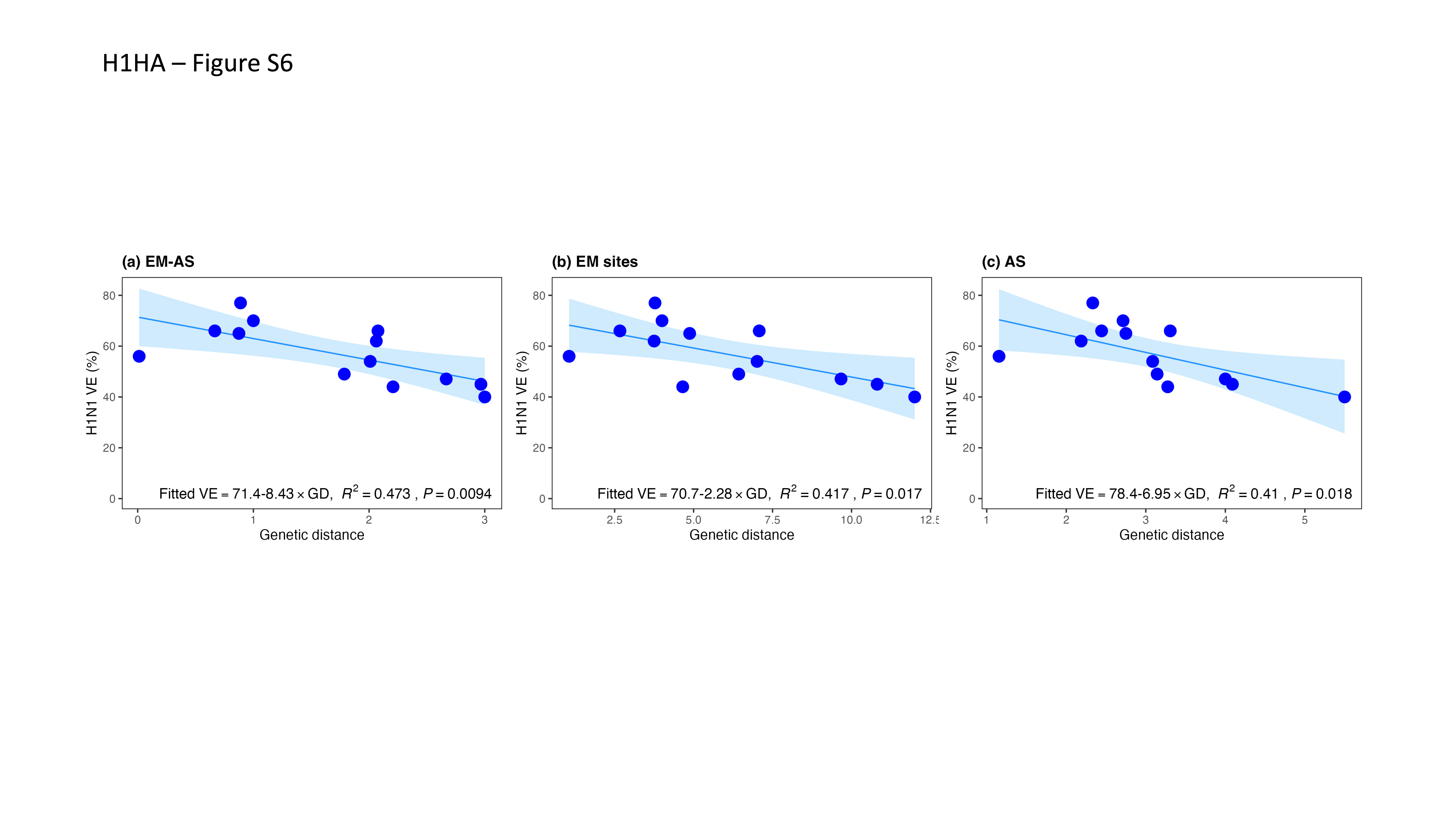

#### Figure S6. Comparing prediction performance of three predictor codon sets of HA for H1N1 virus

(a) H1N1 VE against GD on the intersection set of EM and AS (EM-AS); (b) H1N1 VE against GD on EM sites; (b) H1N1 VE against GD on all AS.

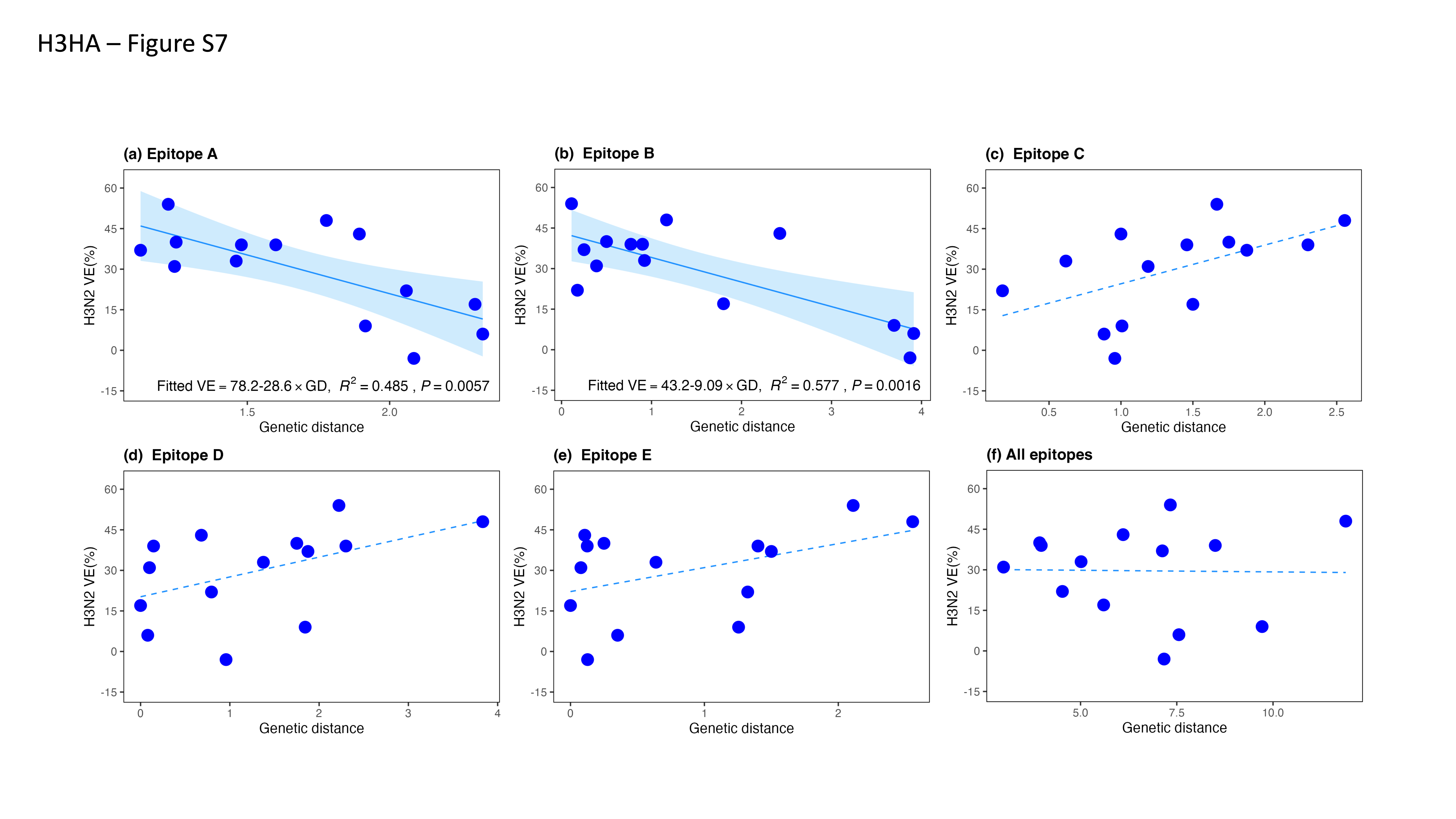

#### Figure S7. The relationship between H3N2 VE and genetic distance on five epitopes of H3N2 HA protein

(a) H3N2 VE against GD on epitope A of HA; (b) H3N2 VE against GD on epitope B; (c) H3N2 VE against GD on epitope C; (d) H3N2 VE against GD on epitope D; (e) H3NA VE against GD on epitope E; (f) H3N2 VE against GD on all epitopes.

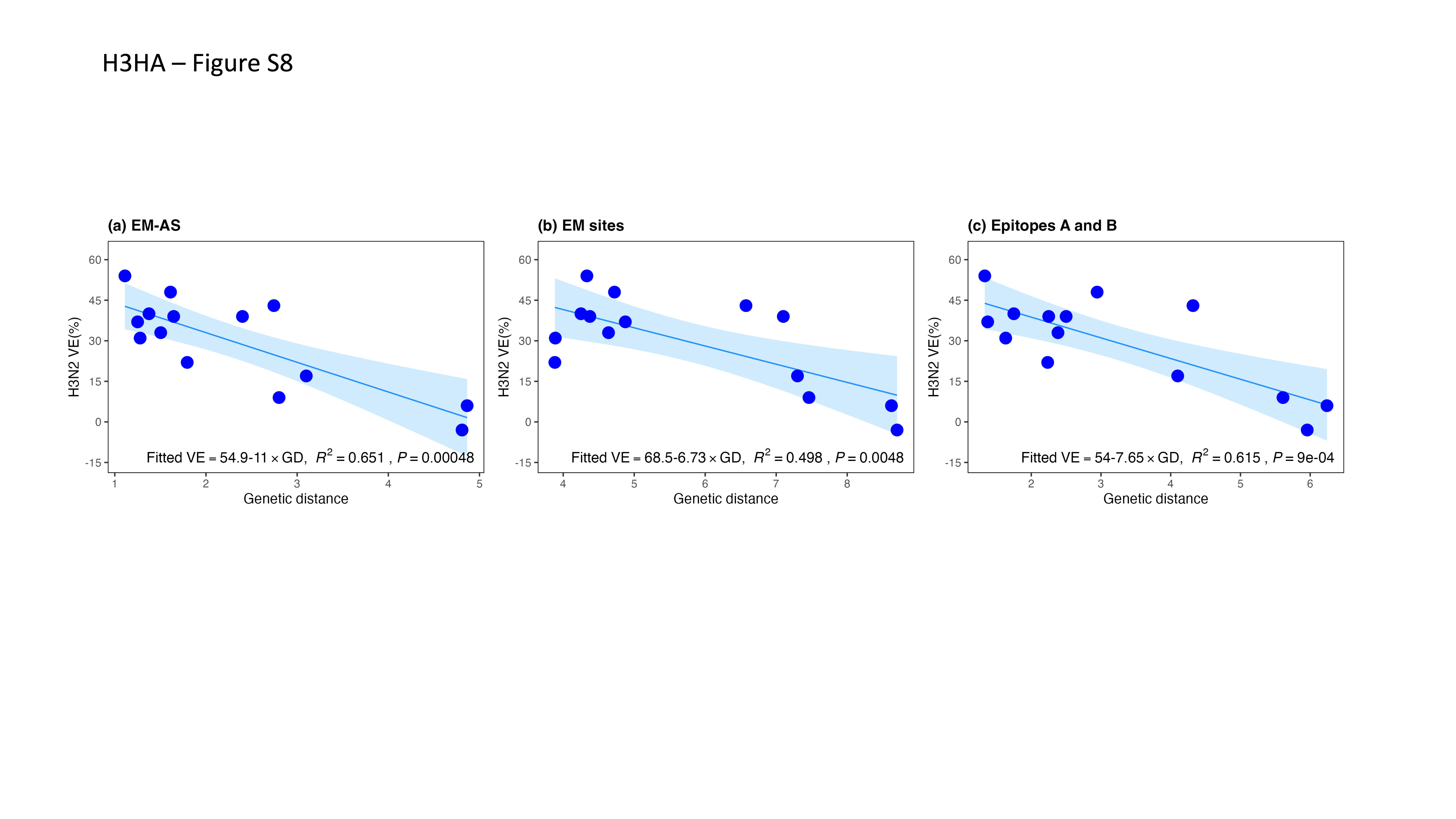

#### Figure S8. Comparing prediction performance of three predictor codon sets of HA for H3N2 virus

(a) H3N2 VE against GD on the EM-AS; (b) H3N2 VE against GD on EM sites; (c) H3N2 VE against GD on epitopes A and B.

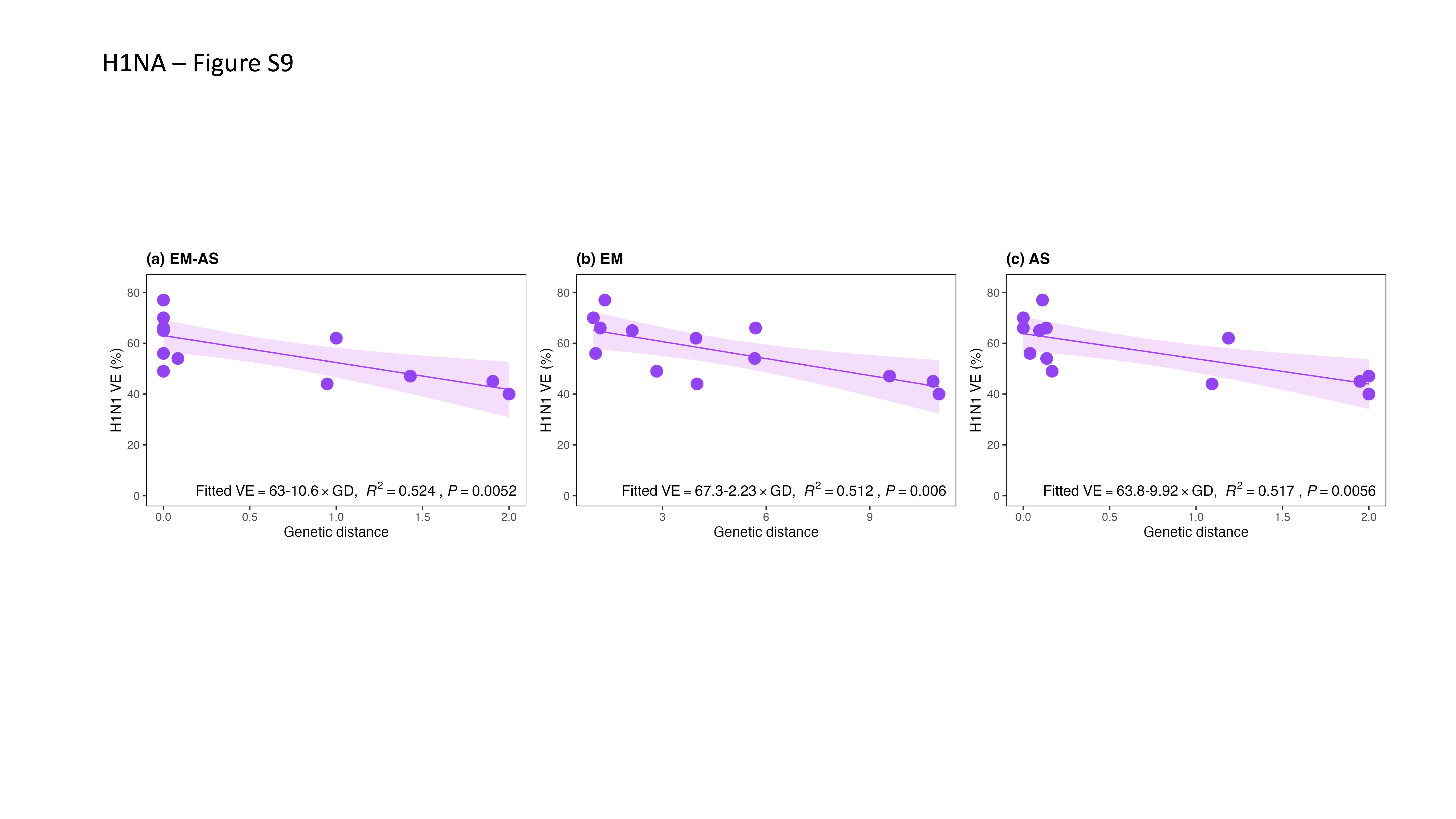

#### Figure S9. Comparing prediction performance of three predictor codon sets of NA for H1N1 virus

(a) H1N1 VE against GD on the EM-AS; (b) H1N1 VE against GD on EM sites; (c) H1N1 VE against GD on all AS.

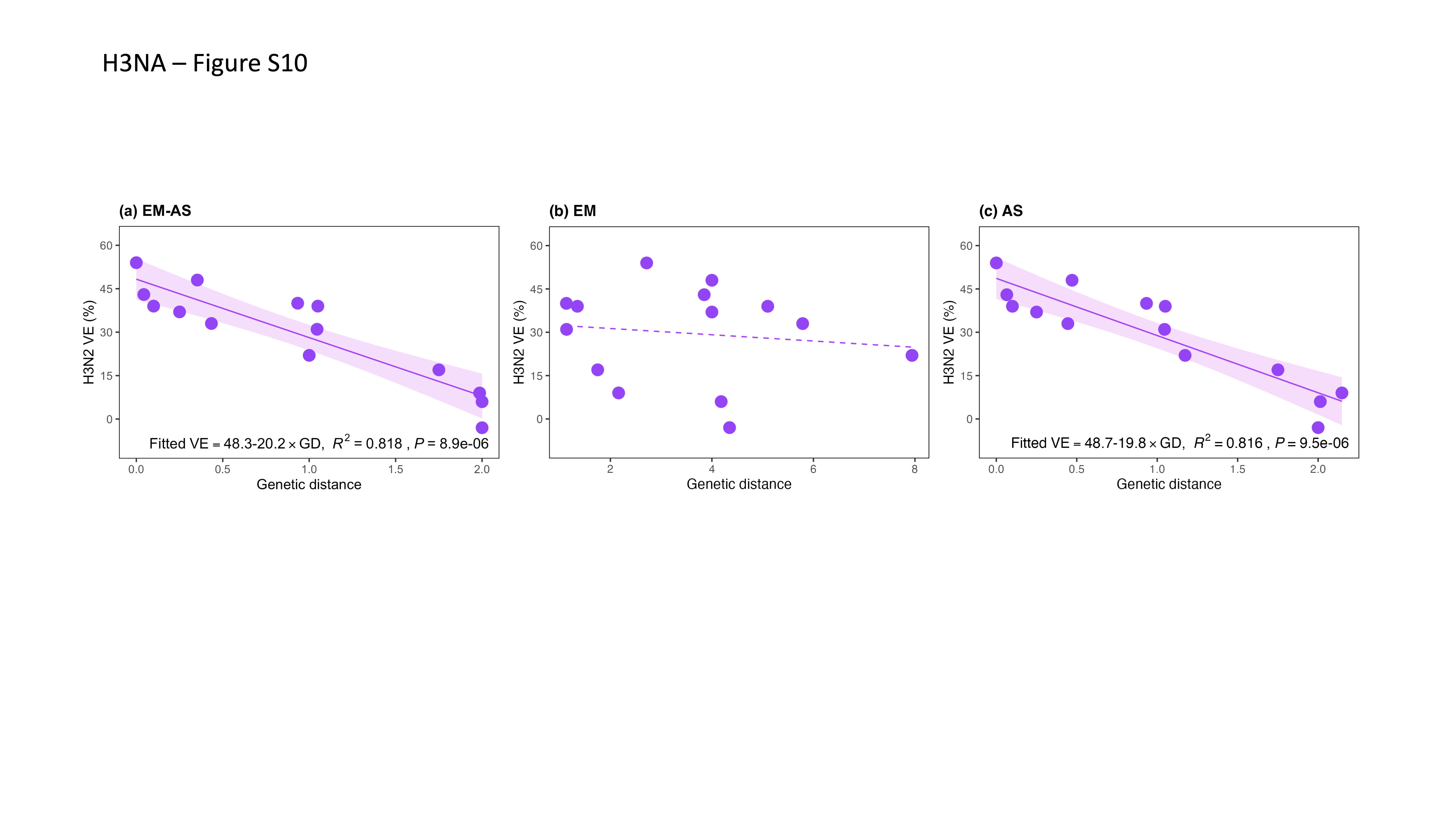

#### Figure S10. Comparing prediction performance of three predictor codon sets of NA for H3N2 virus

(a) H3N2 VE against GD on the EM-AS; (b) H3N2 VE against GD on EM sites; (c) H3N2 VE against GD on all AS.

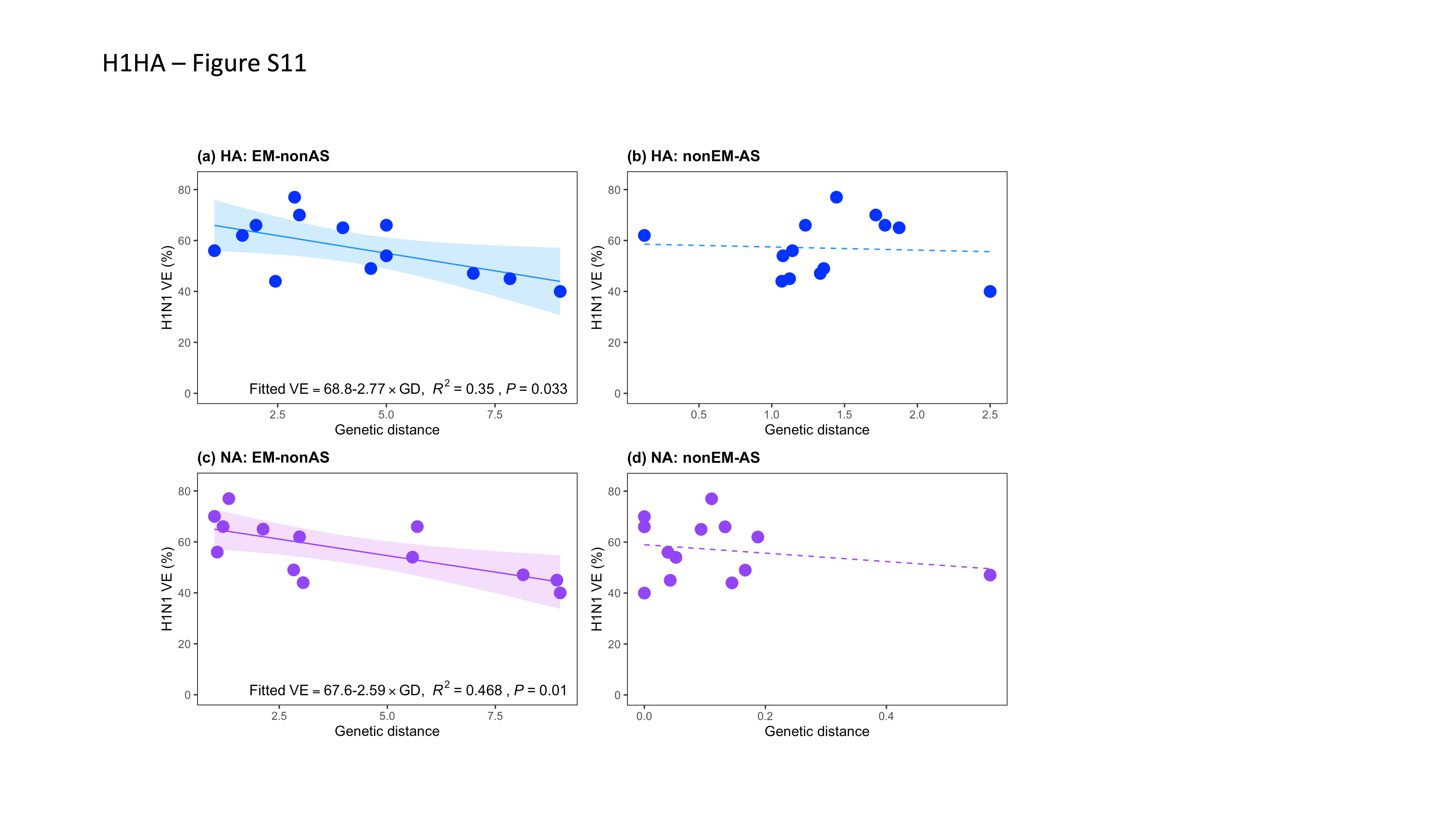

#### Figure S11. The relationship between H1N1 VE and GD on EM-nonAS and nonEM-AS

Panels (a-b) are about HA protein and panels (c-d) are based on NA protein. EM-nonAS means non-intersection subsets of EM to AS and nonEM-AS denotes a locus that is AS but not EM.

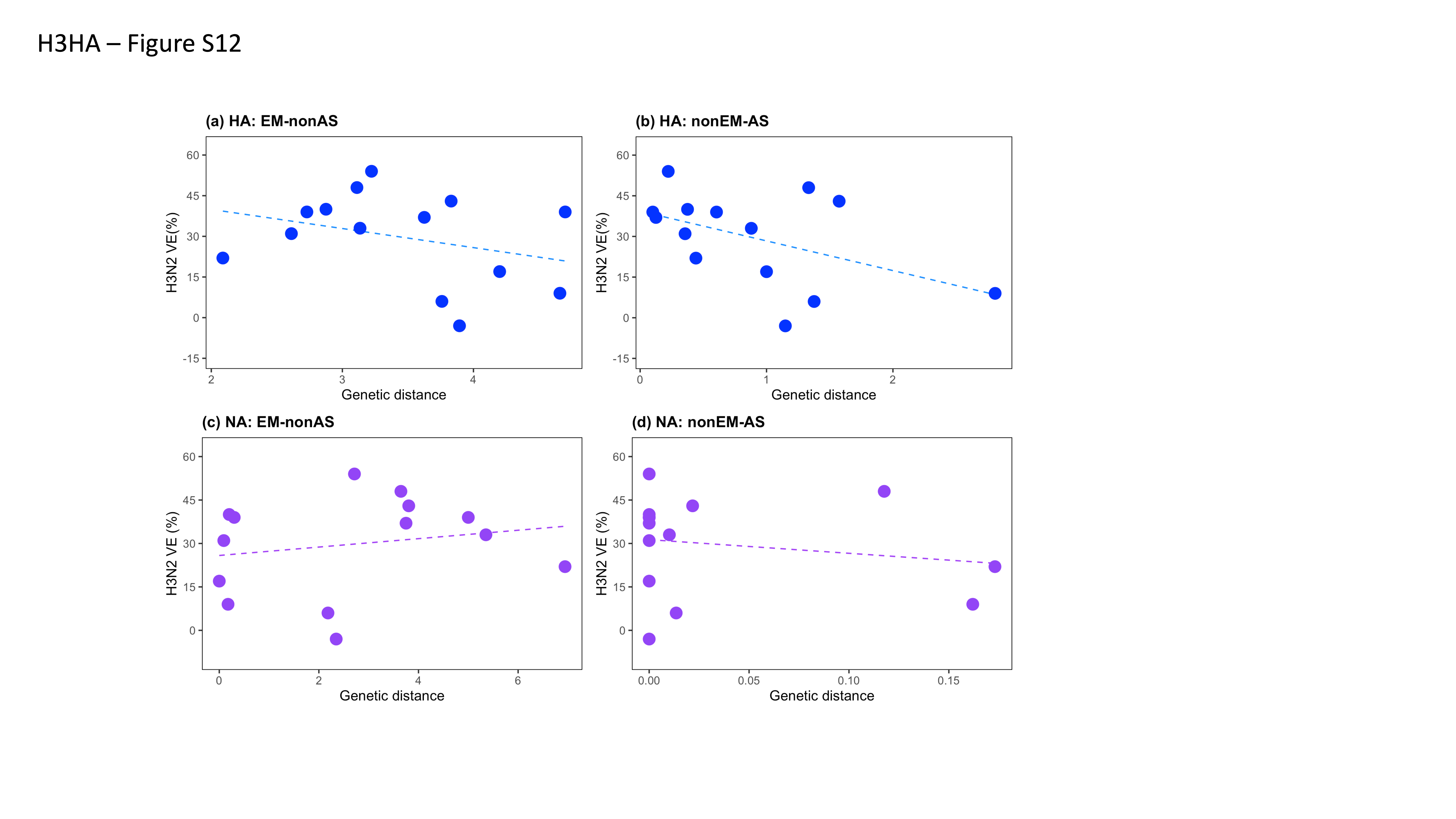

#### Figure S12. The relationship between H3N2 VE and GD on EM-nonAS and nonEM-AS

Panels (a-b) are about HA protein and panels (c-d) are based on NA protein. EM-nonAS means non-intersection subsets of EM to AS and nonEM-AS denotes a locus that is AS but not EM.
